# Evidence on the use of Birthrate Plus® to guide safe staffing in maternity services – a systematic scoping review

**DOI:** 10.1101/2023.10.17.23297132

**Authors:** Peter Griffiths, Lesley Turner, Jenny Lown, Julia Sanders

## Abstract

**Background:** Birthrate Plus® is a widely used tool that informs decisions about the number of midwifery staff needed to provide safe and high quality care in maternity services.

Evidence about the effectiveness, validity, reliability, and feasibility of tools such as this is needed.

**Objective:** To identify, describe and analyse the available evidence supporting the use of Birthrate Plus.

**Methods:** We searched PubMed, Medline, CINAHL, Google Scholar, Scopus, Academic Search, British Library Ethos, Directory of Open Access Journals and Science Direct. Studies were eligible if they reported empirical data relevant to the validity, reliability, or useability of Birthrate Plus or if they measured the impact on staffing levels, outcomes, costs or provided a comparison with other methods.

**Results:** 23 sources of evidence were identified and reviewed. We found no prospective intervention studies on the use of Birthrate Plus to demonstrate outcomes for mothers, babies or staff wellbeing. Nor did we find studies comparing the tool to other methods or addressing resource use. Most of the evidence was descriptive, focussing on the use of the tool or the results of Birthrate Plus assessments. There is some evidence of the reliability of application of categories within the tool, the ability of the tool to detect variation in demand and to highlight staff shortages.

**Conclusions:** In terms of traditional hierarchies of evidence, the evidence for Birthrate Plus is weak. There is a need for more independent research or simulation using real world data to understand how the tool performs in the current context of midwifery practice.

**Statement of significance:** *Problem or Issue:* It is important to ensure that there are sufficient midwives to provide safe and effective care and support positive experiences for women during pregnancy and child birth.

*What is Already Known:* Birthrate Plus is a widely used tool to calculate staffing requirements, which is promoted by the company as being ‘evidence based’.

*What this Paper Adds:* This review of evidence found major gaps. There is no direct evidence that Birthrate Plus calculates the correct level of staffing or performs better than other systems or professional judgement alone.

## Background

Maintaining safety, the quality of care and staff wellbeing in maternity services requires that there are sufficient midwives to meet need in all areas of the service including ante-natal, delivery and post-natal care. The Birthrate Plus® (BR+) staffing tool is designed to assess the clinical care needs within maternity services, and to use this data to determine the number of midwives employed to meet this need consistently.[1] The tool seeks to ensure that sufficient staff are available to deliver one to one care during labour and meet other needs for antenatal and post-natal care. BR+ is widely used throughout the UK National Health Service and has also been used in Australia, China, Ireland and the Netherlands. [2–4]

It is essential that any tool that is widely used can determine the correct number of staff, ensuring that there are sufficient staff to meet predictable variation in demand while maintaining standards without wasting scare resources. There is evidence of harm if maternity services are understaffed. Adverse events such as increased rates of perineal damage, postpartum haemorrhage, maternal readmission, and neonatal resuscitation are associated with low midwifery staffing.[5] Investigations into healthcare failures have scrutinised staffing levels.[6, 7] In the UK, care failings in one NHS Trust led to an investigation, the Ockenden report[8], which highlighted the need to staff units appropriately for the complexities of 21^st^ century maternity care, ensuring there is sufficient staff in the right place at the right time and to agree minimum staffing levels. The report found that that numbers of midwives available on each shift were frequently insufficient to provide safe care. Increasingly, there is also concern that inadequate staffing levels impacts negatively on the wellbeing and retention of midwives.[9]

Although midwife staffing levels are not prescribed or legislated for in most countries, evidence has associated continuous support in labour with better outcomes. Outcomes include improving childbirth experience, increasing spontaneous vaginal birth, reduced duration of labour and need for pharmacological analgesia and fewer babies with low Apgar scores.[10] In the United Kingdom one to one midwifery care during labour has been the expected standard of care for the National Health Service since 1980.[11, 12] This standard was incorporated into the intrapartum midwife time calculations within the Birthrate tool (subsequently Birthrate+), when developed in the mid 1980’s. Similar standards of one to one care during labour are the basis of staffing standards for maternity case elsewhere, including the USA.[13]

At the core of BR+ is an assessment which is used to classify each intrapartum episode and to indicate the relative staff time required. The assessment is multi-factorial with sections addressing gestation and labour, delivery, infant(s) and intensive care factors.[14] Starting with a requirement for one to one care for the duration of labour, the midwifery time requirements are inflated above one to one for those with more complex care needs, similar to staffing models in intensive care units.[15] The tool also calculates the hours of work associated with admissions without delivery and required midwifery time to provide antenatal and postnatal care. Additional allowances are made for non-clinical work, leave and sickness before converting the calculations into the Full Time Equivalent staff required.[14] Material supporting the use of Birthrate Plus emphasises that the tool should be used alongside professional judgement, including recognition of individual service need, to set establishment levels.[1]

Birthrate Plus is widely used and made available as a paid service by a UK based company.[16] Birthrate Plus is described by its developers as ‘validated’ and ‘evidence based’[16] and publications relating to Birthrate Plus span over 30 years. However, the evidence base has not previously been systematically evaluated. A review of evidence for decision support tools for the National Institute for Health and Care Excellence[17] excluded most published accounts of evidence about the development of Birthrate Plus because they were descriptive and provided no evidence that Birthrate Plus resulted in changes in staffing or outcome. The developers have published a number of papers outlining current methods and its application in maternity settings, but details of the tools’ development and underpinning assumptions are sparse.[1] [14] [18–20] They report that formulas have been updated and extended over time as the methodology has developed[14], but such changes remain unpublished. A version of the tool has been developed to monitor fluctuations in workload and midwife availability in real time via an App.[19] In 2014, The National Institute for Health and Care Excellence endorsed Birthrate Plus as a tool includes most of factors highlighted their safe staffing guideline[21], however they also made clear that this endorsement was not based on a judgement about the quality of the tool, a direct assessment of evidence or tests of the decision algorithms within it.[22]

The Ockenden report into care failings in one UK maternity service[8] called for the feasibility and accuracy of the Birthrate Plus tool to be evaluated. Because of the widespread use of the tool and the claims made about it, it is timely to review the evidence more broadly to understand what is and what is not known about the performance and utility of Birthrate Plus.

## Methods

We undertook a scoping review in order to identify and map the available evidence.[23] Because early searches identified limited evidence and because a key aim of our scoping review is to describe the existing evidence base, we included all sources that reported empirical evidence relating to the Birthrate plus system. While we were open to consider any evidence, we considered this evidence within a framework based on questions and issues highlighted in reviews of evidence for nurse staffing methods and used these questions to organise reporting of study results.[24]

1. What evidence is there for the reliability and validity of Birthrate Plus assessments or the resulting establishment estimates?
2. What evidence is there for the useability / perceived usefulness of Birthrate Plus in workforce planning?
3. What evidence is there for the impact of Birthrate Plus based establishments on planned / achieved staffing?
4. What evidence is there that Birthrate Plus addresses variability in demand within services (from day to day and hour to hour) and across settings
5. What evidence is there for the impact of Birthrate Plus on quality of care / outcomes for parents / babies?
6. What are the costs and / or cost effectiveness of using Birthrate Plus?
7. How does Birthrate Plus compare with other methods to determine staffing in maternity services?

We searched British Library Ethos, CINAHL+, The Cochrane Library, Ebsco OA database, Google Scholar, PubMed (Medline), Science Direct, Scopus and Web of Science. Consistent with a scoping review we developed our search strategy iteratively. Preliminary searches used a complex structured search including broad terms related to staffing tools (e.g. Birthrate Plus OR Acuity tool OR Workforce Planning OR Staffing Needs) and midwifery staffing (e.g. Midwifery workforce OR Maternity staffing OR Establishment OR staff Planning OR Midwifery Staffing). We also searched the Birthrate Plus website and retrieved sources identified as evidence by the Birthrate Plus team.[25–27] We scrutinised the reference lists of relevant material identified at this stage to identify further sources of evidence. It became clear that all relevant studies named the tool, so a more specific and sensitive approach to searching was adopted, with final searches of all databases undertaken in April 2023 simply using the term “Birthrate Plus” (searching in title, abstract and all other fields, including full text if available). No limitation by language, date of publication or country of origin was applied.

We included all sources in peer reviewed journals that provided some empirical evidence, excluding only general reviews and discussion pieces, although reference lists of such papers were scrutinised. Empirical research from non-peer reviewed sources was eligible for inclusion and because of the potential significance of sources described by the tools owners as ‘evidence’ we included all sources cited as ‘evidence’ on the Birthrate Plus website. Eligibility for inclusion was assessed by two reviewers. Search results and decisions at each stage of screening are summarised in a ‘Prisma’ flow chart (Figure 1).

**Figure 1.**
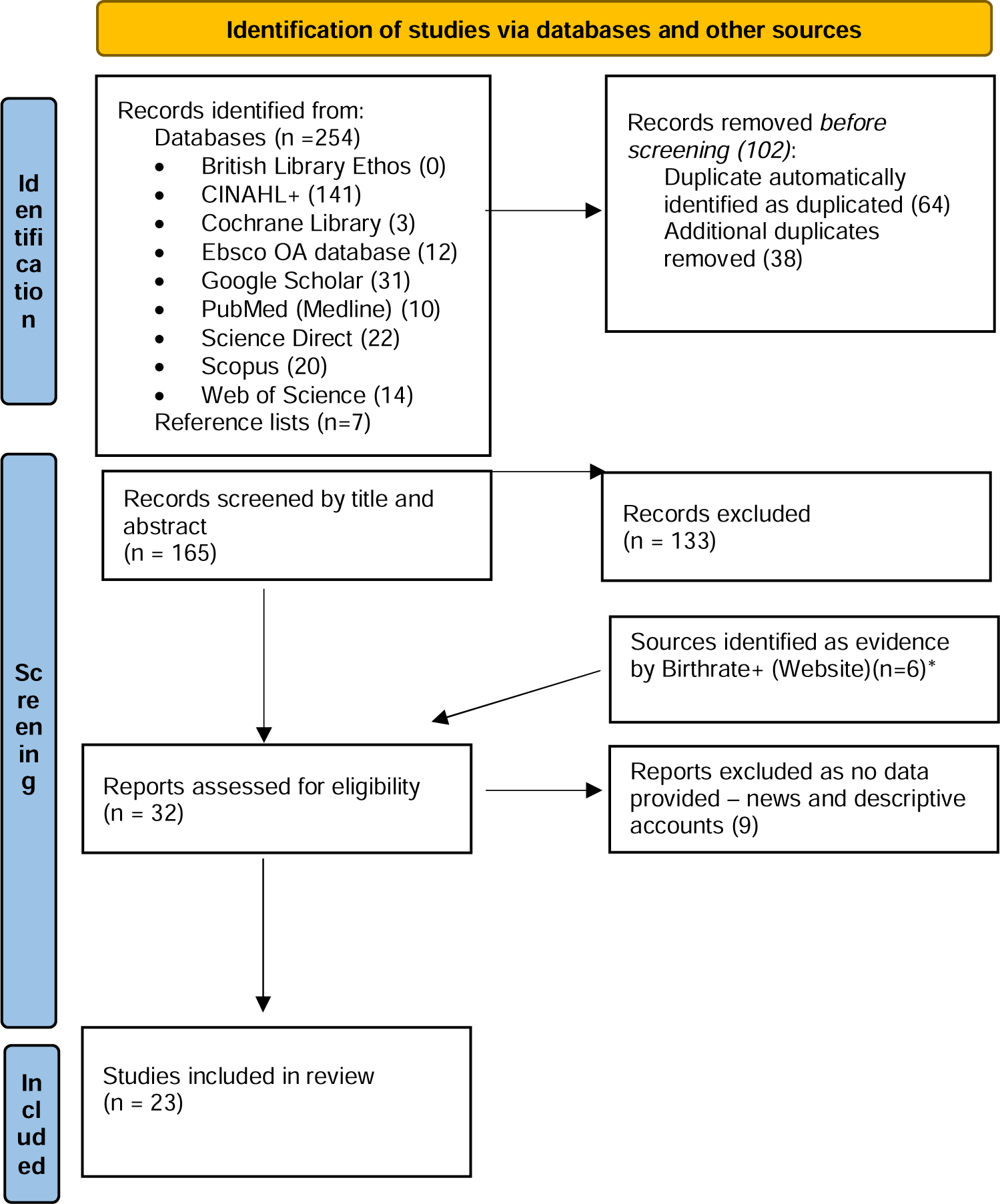
PRISMA flow diagram. *From:* Page MJ, McKenzie JE, Bossuyt PM, Boutron I, Hoffmann TC, Mulrow CD, et al. The PRISMA 2020 statement: an updated guideline for reporting systematic reviews. BMJ 2021;372:n71. doi: 10.1136/bmj.n71 *Sources identified by Birthrate Plus as ‘evidence’ were included in the review although they may have otherwise been screened out at other stages if they included no empirical data

Data were extracted by one reviewer and charted to include authors, publication date, study design, setting, findings and which of the seven study questions were addressed by each paper. Findings were synthesised for each study question. In keeping with the scoping review methodology, we did not undertake a formal critical appraisal of individual studies as the questions they addressed were diverse, as were our aims in reviewing the evidence. However, we considered the quality of evidence in more general terms in relation to hierarchies of evidence for questions of accuracy and effectiveness. As this scoping review developed iteratively based on leads available within the literature and evidence supplied by Birthrate Plus, we did not register a protocol in advance.

## Results

We identified and accessed 23 sources of evidence for review, of which 15 were published in peer reviewed journals. Of these, two were available only as abstracts (See Table 2). Four sources were published before 2000, 12 were published between 2000 and 2014 and 7 published from 2015 to 2023. Seventeen publications in total reported some empirical data although this was based on the results of Birthrate Plus assessments undertaken for workforce planning, as opposed to being designed primarily to evaluate the tool. In some cases, use of Birthrate Plus was somewhat incidental and any inference made about the tool is indirect. Fourteen studies could be characterised as observational descriptive studies while three were more analytical, using simulation models. Eleven sources were co-authored by Ball, who developed Birthrate Plus. Of these, six provided some descriptive empirical data arising from the use of the tool[1] [19] [26] [28–30]. We found no prospective intervention studies using Birthrate Plus.

**Table 1.** Search Strategy.

|  |  |  |  |  |
| --- | --- | --- | --- | --- |
| Maternity | **AND** | Birth-rate Plus® | **AND** | Reliability |
| Midwifery |  | BR+ |  | Validity |
| Mid* |  | Staffing ratio |  | Safe staffing |
|  |  | Workload |  | Accuracy |
|  |  | Personnel Staffing |  | Acuity |
|  |  | Scheduling |  | Experience |
|  |  | Workforce |  | Evaluation |
|  |  | Staff planning |  |  |
|  |  | Establishment |  |  |

**Table 2:**
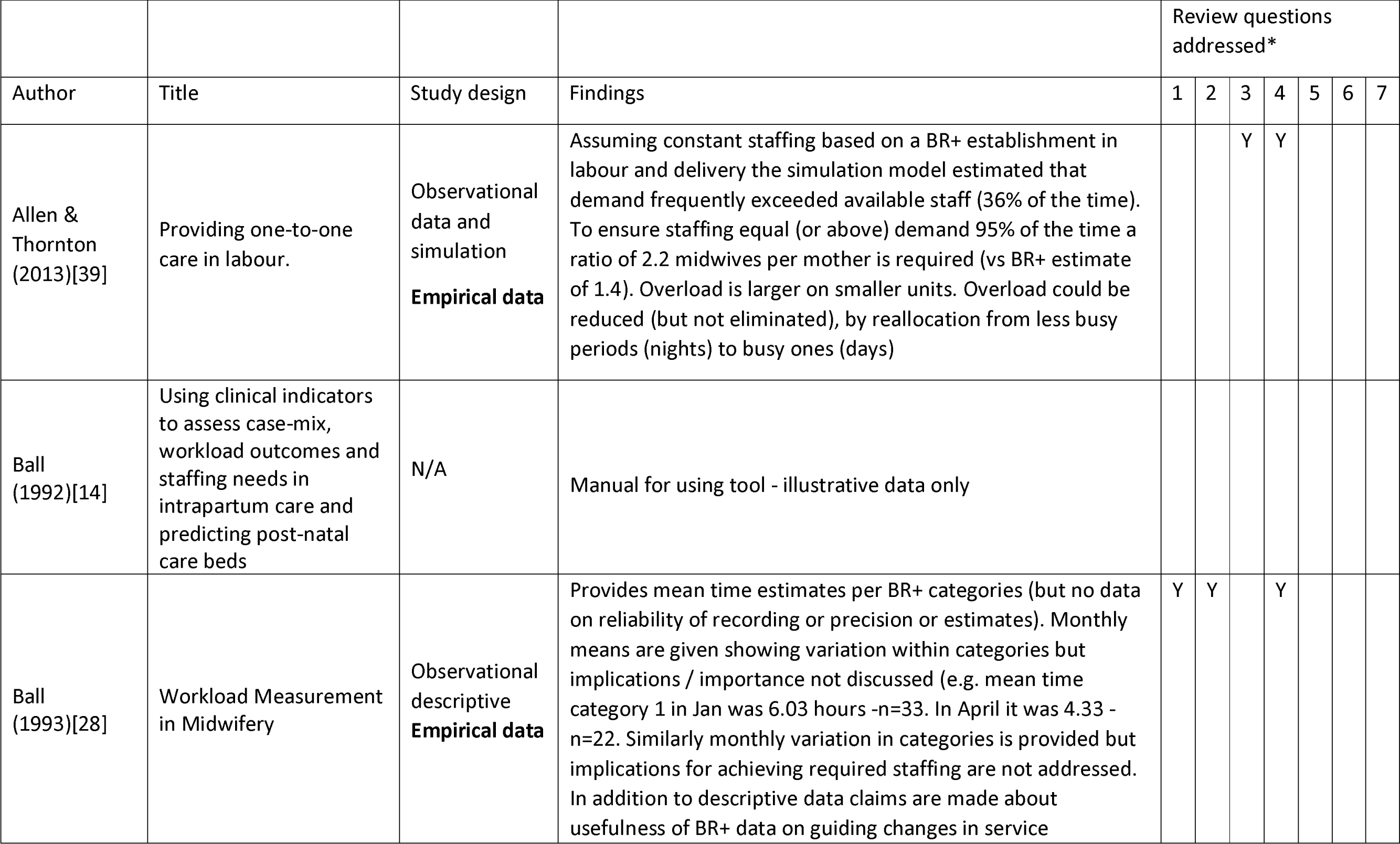
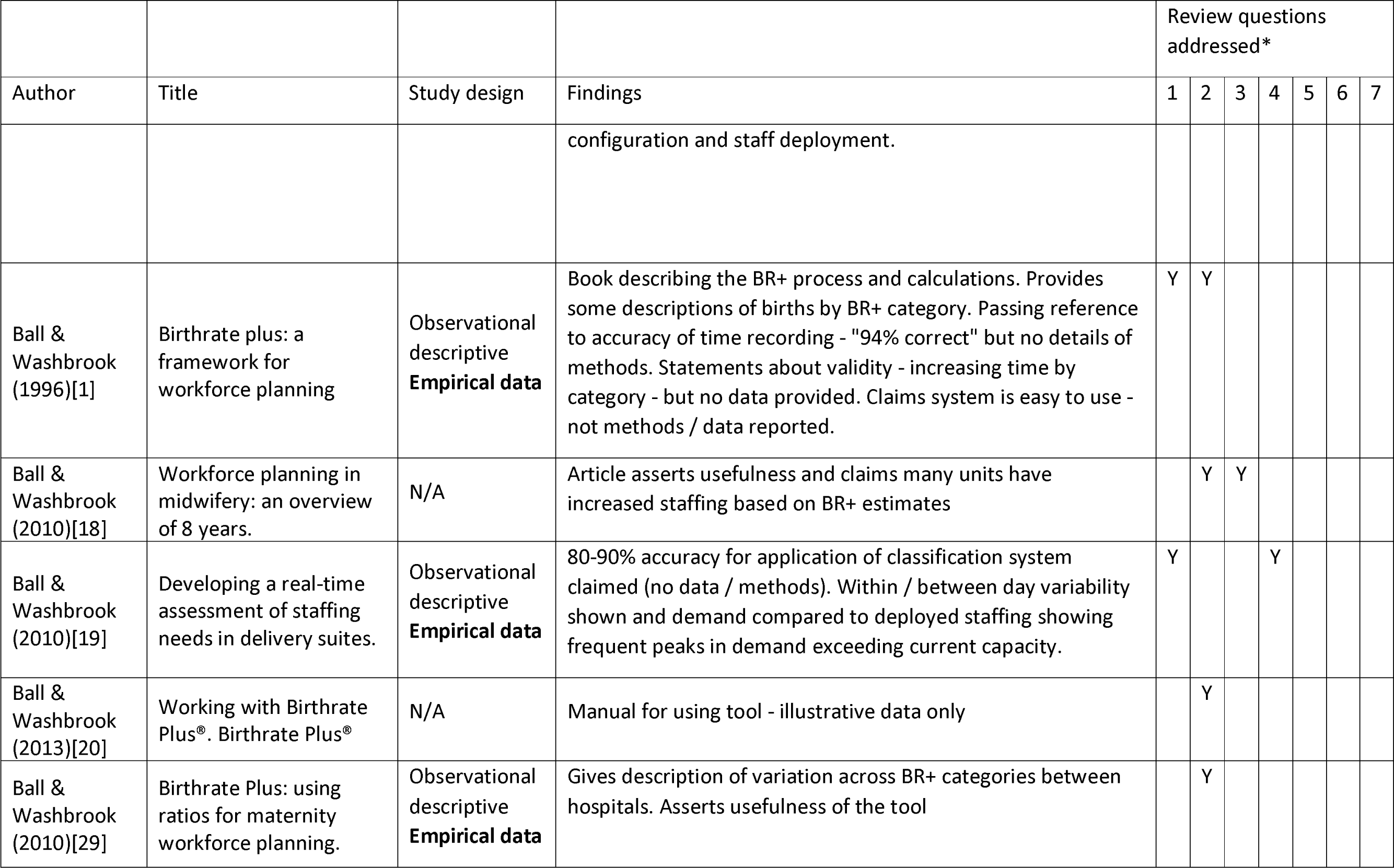
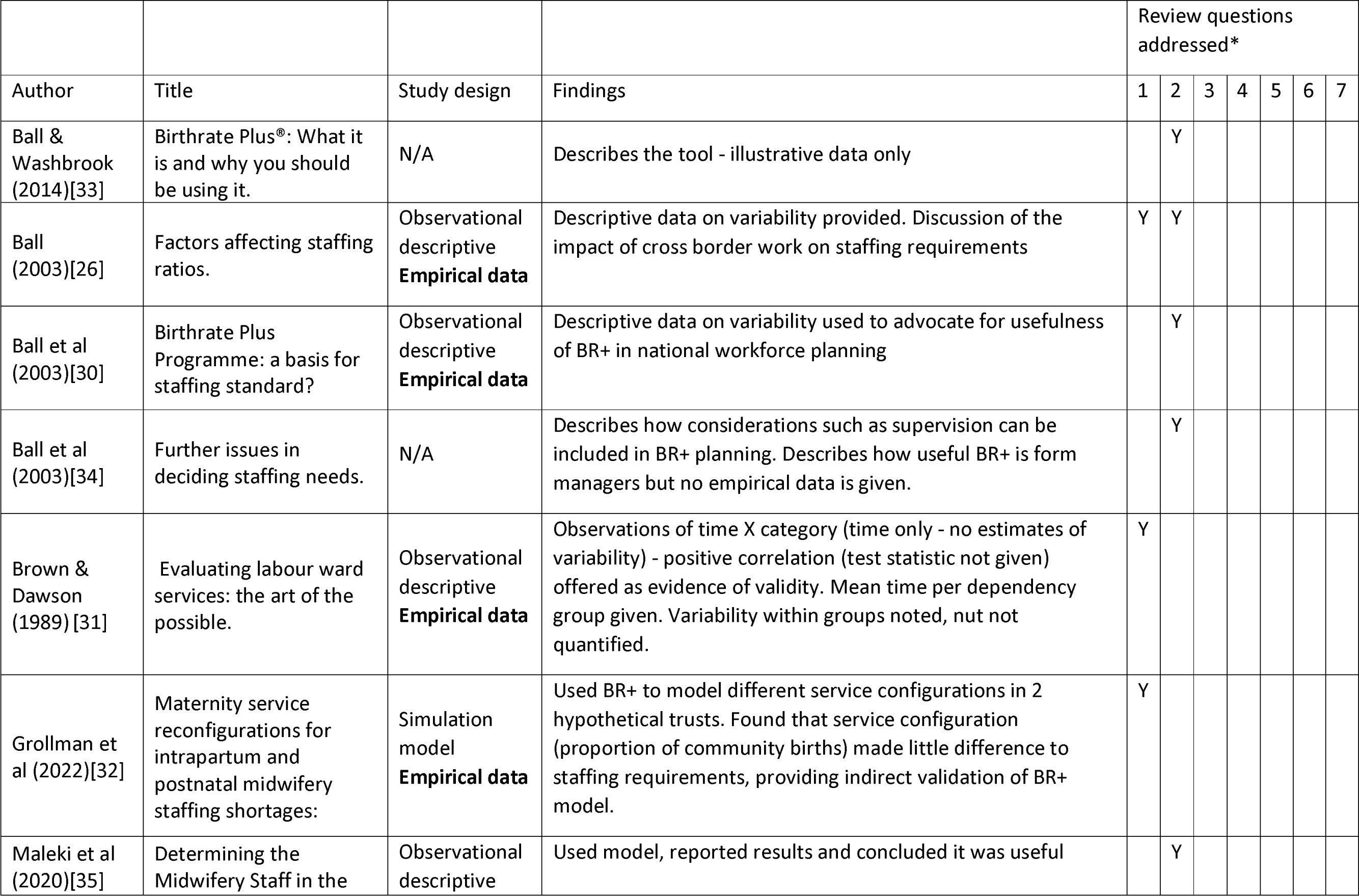
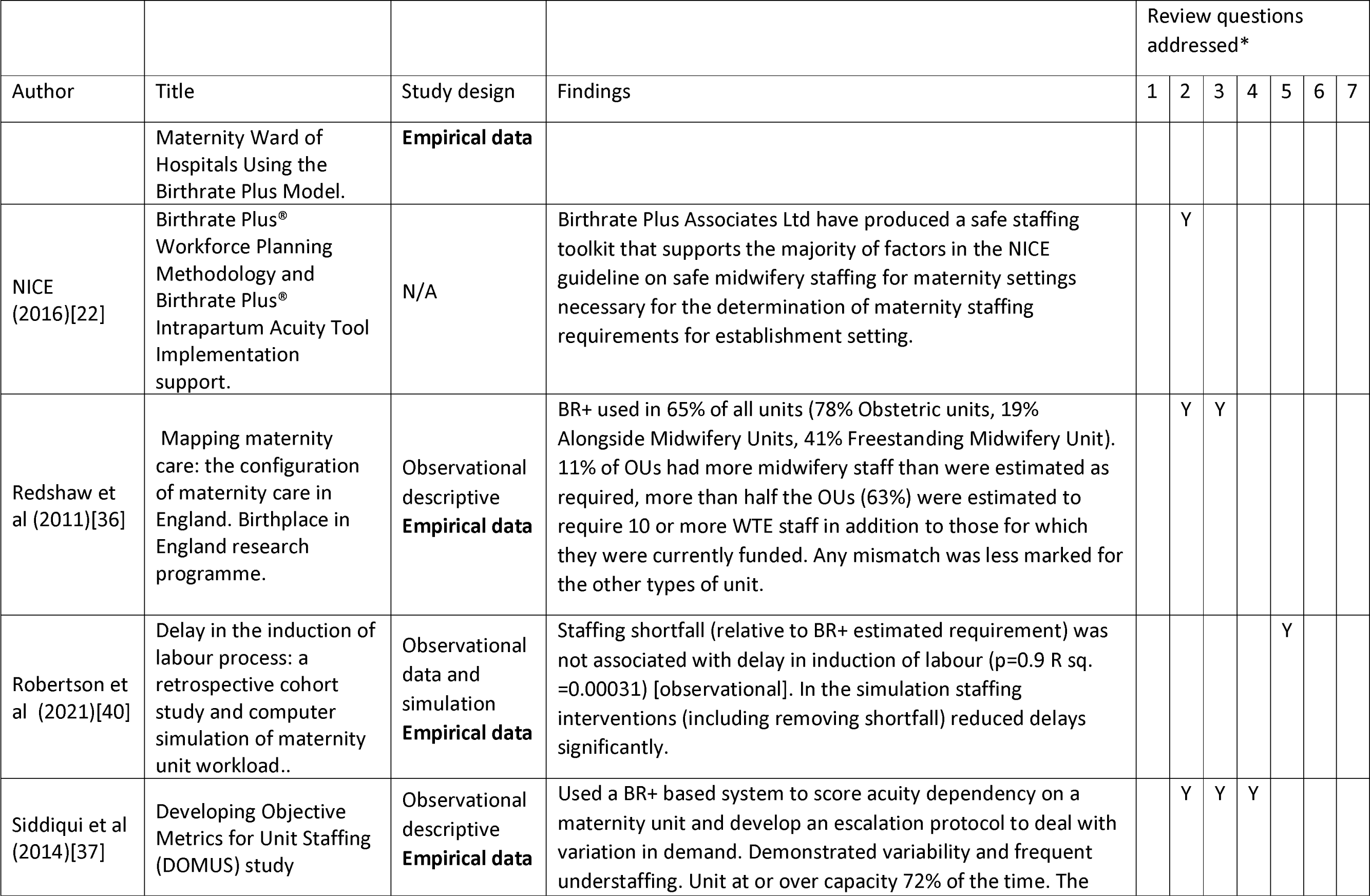
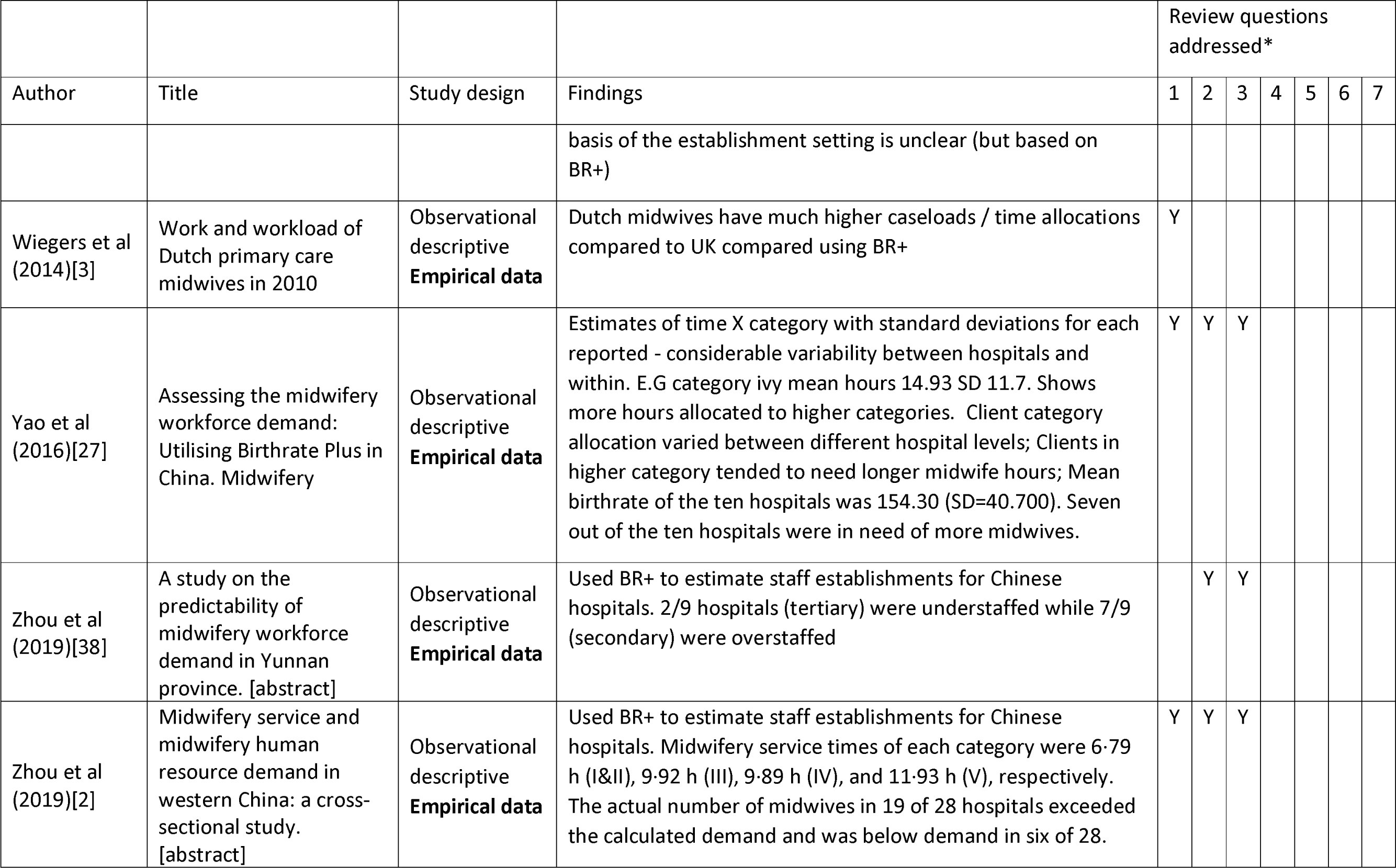
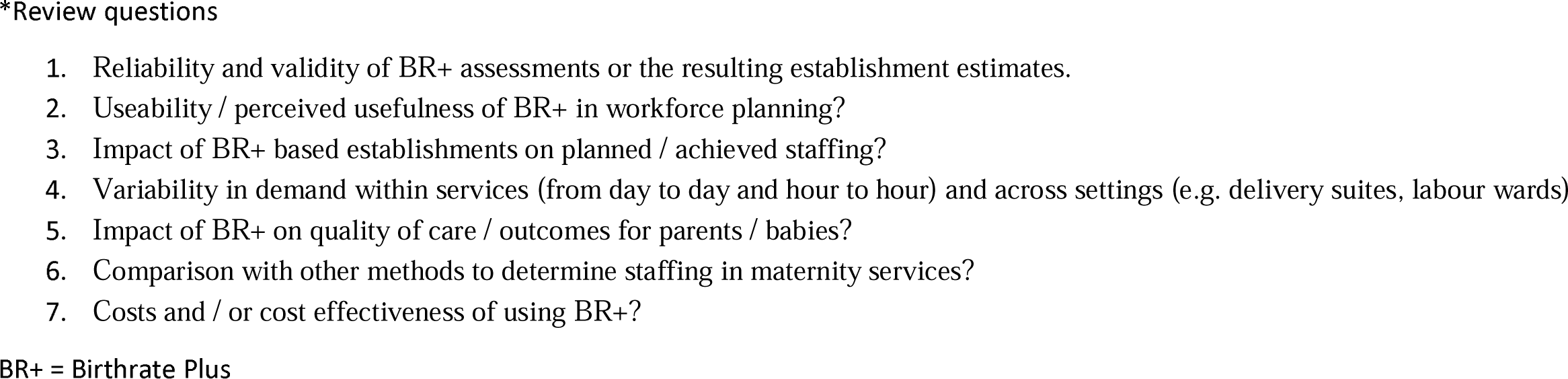
Evidence table.

While we do not offer a detailed methodological critique of the studies summarised below, none would be considered as providing robust evidence of the benefits or accuracy of Birthrate Plus using traditional hierarchies of evidence. There are no controlled trials providing evidence of effectiveness in terms of outcomes for mothers, babies, or staff and no comparisons of Birthrate Plus based staffing estimates with any clear criterion, certainly nothing that might be considered a “gold standard”. Many of the inferences made are indirect and many of the empirical claims made in sources are based on imprecisely described methods.

### Reliability, validity & comparison

Nine sources were judged to offer some evidence of reliability and validity of the tool, although none compared results of the Birthrate Plus assessment with others. [1–3] [19] [26–28] [31, 32] Claims for reliability were based on reference to accuracy of midwives recording of the time women were on delivery suite compared to that recorded in clinical notes (94% correct)[1] and classifications (80-90% accuracy for selection of intrapartum complexities)[19] but without details of the methods used for this assessment. Several sources offer support for the validity of the Birthrate Plus classifications by showing increasing midwifery time was required to provide care to women with labour complexities. [2] [27–29] However, methods used to assess the original time ratios are not available and the continued accuracy or relevance of the time estimates have not been reviewed since the original development. One source refers to considerable variability of time within categories[26] while one study of Chinese maternity units shows very large standard deviations on time estimates. [27] For example for category I births without any complexity or need for medical involvement, the mean time was 6.4 hours with a standard deviation of 5.2 hours. Other reports show considerable variation in case mix from month to month within units.[28] The implications of this for the accuracy and precision of establishment estimates are not clear, although guidance for the use of Birthrate Plus recommends that large samples and prolonged periods of data collection are needed to establish midwifery staff requirements in any unit.

Indirect evidence comes from two studies. A simulation study that used Birthrate Plus methods in a model of different service configurations in two hypothetical trusts found that the proportion of planned community births made little difference to staffing requirements. [32] A study of Dutch midwifery services found that time allocations and caseloads inferred using the Birthrate Plus classifications were dramatically different (50% or more) from current practice in the Netherlands.[3] The direct relevance of this is unclear other than to raise questions about whether the assumed staffing levels in the model translate into practice, the generalisability of the method, and the relevance of the underlying staffing standard when other aspects of the health system change.[3]

### Useability, Perceived Usefulness of Birthrate Plus in workforce planning

While 16 of the 23 studies were assessed as offering some evidence of the useability of the tool, the evidence was almost exclusively based on the conclusions or assertions of authors rather than direct empirical evidence. [1, 2] [18] [20, 22, 26-30] [33–38] No empirical evidence about staff experiences of using the system, or the time involved, was found, although several reports referred to the system’s usefulness and ease of use. [18, 20, 28, 34, 35] There was no empirical evidence that staff felt the recommended staffing levels were appropriate. The NICE statement of endorsement did not provide any empirical evidence but determined that the tools (both for establishment setting and the real-time acuity assessment) addressed many of the factors needed in setting maternity staff establishments.[22]

One UK study used a tool based on Birthrate Plus to measure real-time demand and used results to guide an escalation protocol to deal with variation in demand on labour ward, over a 12-month period.[37] There was considerable variability in demand and frequent understaffing, with the unit operating at or over capacity 72% of the time, although critical understaffing (large or persistent staff shortages) occurred on only 14 occasions. Authors conclude that real-time monitoring using a classification based on Birthrate Plus was useful. We found no studies which compared the usability or application of Birthrate Plus to other staffing tools or methodologies and no studies that explored the resource use or costs of applying the system.

### Impact of BR+ based establishments on planned / achieved staffing

Birthrate Plus is said to have usefully guided changes in staffing in response to change in service delivery[28] and led to increased staffing.[18] Birthrate Plus is widely used[36] and several studies showed estimated staffing requirements that differed from existing levels and so would have led to change if implemented. [2, 36, 38] Birthrate Plus has been used in simulation studies to compare establishment staffing with demand[39] and has also been used to underpin escalation tools for delivery suite prompting an operational response to maintain appropriate staffing levels.[37] We found no studies that reported the costs of staffing changes associated with Birthrate Plus.

### Ability to identify variability in demand

Several studies provided evidence of variability in demand within days and from day to day.[19] [37] [39] To accommodate variability and peaks in demand, Birthrate Plus allows 15% reserve resource when estimating required establishments[39]. Two studies showed demand frequently exceeding the capacity of available staff[19, 37], but it was unclear if the establishments had been set using Birthrate Plus (although it seems likely), or whether available staffing levels matched those recommended. Thus, inferences about the ability of Birthrate Plus based establishments to meet variable demand cannot be made from these studies. Assuming constant staffing based on a Birthrate Plus establishment in labour and delivery, a simulation model estimated that demand exceeded available staff 36% of the time.[39] To ensure staffing equal (or above) demand 95% of the time a ratio of 2.2 midwives per mother was required, which contrasts with the range of Birthrate Plus estimates of 1.0 to 1.4 midwives per mother. Overload could be reduced (but not eliminated), by reallocation of staff from less busy periods (nights) to busy ones (days). [39]

### Impact of BR+ on quality of care / outcomes for parents / babies

A single UK study addressed the impact of staffing below the Birthrate Plus recommended levels on delays in induction of labour. Delays were associated with high numbers of women booked for induction or presenting in spontaneous labour. Observational data showed no association between staffing shortfalls and delays in induction in labour, although a simulation model suggested that staffing interventions, including reducing shortfalls would reduce delay.[40] No other evidence of effects on quality of care or outcomes were found. We found no studies that addressed the costs or cost-effectiveness of using Birthrate Plus in relation to any outcome or quality measure or when compared to any ither approach to determining staffing requirements.

## Discussion

We found a large body of published work including peer reviewed papers describing the use of Birthrate Plus. However, the extent of empirical evidence provided is very limited and there has been little scrutiny of the system’s methods or performance. A significant proportion of the peer reviewed sources have been produced by the originators of the system, but most empirical data these accounts provide is purely descriptive with other claims backed by poorly reported data or anecdote. Much of the independent research has incorporated, rather than evaluated Birthrate Plus methods, although results have provided some insights into variability across settings. There is no empirical evidence of the benefits of using Birthrate Plus compared to other approaches to setting establishments.

Key questions when evaluating staffing tools are whether they lead to altered staffing levels, improve the quality of care for women, improve staff wellbeing and retention, and whether the estimated staffing requirements are in accord with professional judgement.[41] The consequences of understaffing are known in terms of poor care experiences, outcomes, morale and retention of staff.[5, 42] A common reason for midwives leaving the profession is that they are not able to give the quality of care that is needed, that staffing is unsafe or they are experiencing burnout.[43] There is sufficient evidence of associations between midwifery staffing levels and outcomes to infer that any increases in staffing levels guided by Birthrate Plus are likely to be associated with improved quality of care.[5] However, while there is evidence that Birthrate Plus has been widely applied, it is less clear whether its use has resulted in changes in staffing plans and whether changes have reduced staffing shortfalls. There is no evidence that the calculations produced by Birthrate Plus give the optimal staffing levels (irrespective of the criteria used) or perform better than other systems or professional judgement alone.

There is some evidence that staff can accurately and reliably record Birthrate Plus categories and this may have increased with the use of data captured in electronic maternity information systems, although improved accuracy depends on the accuracy of the underlying record. There is also evidence that the incremental categories in Birthrate Plus reflect the increase in the average time needed by women with complexity, which lends some support to the validity of the tool. On the other hand, there is evidence that demand within categories could be highly variable and it is unclear if the Birthrate Plus recommended staffing levels are optimal in terms of meeting variable demand. The assumption that an establishment based on average time plus 15% is sufficient has not been tested but the simulation models indicate that higher staffing than indicated by Birthrate Plus may be needed to avoid frequent shortfalls in the face of variation in case mix and case load. [37, 39] This mirrors the situation for staffing tools in nursing, where scant attention has been given to issues that arise from the variability in the amount of time required within categories or the limited ability of a fixed establishment to meet variable demand. [24]

As used by the evidence-based practice movement the term ‘evidence-based’ implies that the answers to the important questions about the effects, cost-effectiveness or the accuracy of the tool have been answered by robust evidence[44], but this is not the case for Birthrate Plus. Costs and cost effectiveness were not reported in any study. The benefits from any resulting staffing changes need to be measured and judged against the costs, including the costs of any extra staff employed and the costs of using the tool (including payments associated with licensing the system and staff and systems to provide the required data). Staff perceptions and experiences of using the tool were not measured in the evidence, despite reports of ease of use in some papers. [1] [35]

While the lack of independent research on Birthrate Plus is regrettable, the situation mirrors that for widely used tools for setting nursing establishments, where many tools have been developed, adopted and discarded with little evidence.[24] This experience should inform a cautious response to the lack of evidence about Birthrate Plus. Absence of evidence is not evidence that the tool does not work and any de-novo approach will suffer a similar (or greater) evidence deficit. There are few alternatives to this methodology for determining staffing in maternity care (see for example [45–47]). Although we have not formally reviewed the evidence for these tools it is unlikely that there is a substantial evidence base. Similar conclusions about lack of evidence have been made about nursing tools, despite a much larger body of publications. [24] The widespread adoption of Birthrate Plus testifies to some degree of perceived utility and so it is a candidate for more rigorous independent evaluation than has been undertaken so far. While the proprietors of Birthrate Plus cannot be blamed if others have not subjected the tool to rigorous scrutiny, it is important that independent and thorough evaluation is permitted and that commercial interests do not stand in the way of this.

The Ockenden report the found numbers of midwives available on each shift were frequently insufficient to provide safe care[8], but no workforce planning tool can in itself ensure safe staffing levels. Future evaluations of midwifery staffing levels in failing and safe units are needed to explore the contributions of staff availability, recruitment challenges and midwifery staffing budgets in addition to the precision of Birthrate Plus to calculate required staff number. This work should sit within a body of research evaluating the impact of midwifery staffing and skill mix on outcomes for staff and service users.[21] Staffing tools have the potential to contribute to improvements in workforce planning if they are shown to be accurate in their predictions, are used consistently, and staffing levels in practice reflect those recommended. More work is needed in this area to address the wide variation in maternity staffing[48] and to understand decisions which lead to successful matching of supply and demand for services. Nonetheless, even where tools are demonstrated to be accurate, they should not be treated as providing a ‘magical bullet’, in the sense of a definitive answer to the staffing requirements. Rather they are one aspect of establishment setting, which also needs to be informed by consideration of local context, prioritisation and professional judgement.[47]

Further independent research is needed on the validity and precision of Birthrate Plus carried out by independent researchers. A key research question is to measure the effectiveness of Birthrate Plus compared with other decision support methods or professional judgement in recommending safe staffing levels. Assessment of the variable need within intrapartum categories is warranted, as is evaluation of the relevance of the ratios of midwifery time set in the 1980’s against current practice standards. Simulation based on real world data could help inform developments and identify critical issues for consideration and possible adaptation of the tool.

### Strengths and Limitations

We searched extensively across many different sources, including sources of ‘grey’ literature. Consistent with our scoping review methodology, we did not undertake a formal critical appraisal of the evidence we found, but the gaps in the supporting evidence are still apparent, notably the lack of empirical evidence on validity, mother and baby outcomes, cost, cost-effectiveness and impact on staff wellbeing. However, any specific claims based on the results of the research we included need to be interpreted with caution. Inclusion decisions were taken by two reviewers, but some elements of formal data extraction were only undertaken by one. However, while this increases the possibility of some errors all authors have reviewed content, reducing the likelihood of any errors that have a material impact on the review.

## Conclusions

The consequences of getting staffing estimates wrong impacts on the quality and safety of care. Claims that staffing decision support tools are ‘evidence based’ should not be taken at face value without appraisal of evidence of accuracy, effects and costs. While there are many published sources concerning Birth-rate Plus, many in peer reviewed journals, the tool cannot be described as ‘evidence-based’ in the way that the term is typically understood by the evidence-based practice movement.

## Data Availability

All data produced in the present work are contained in the manuscript

